# Phase 2/3 open-label study on NVX-CoV-2601 (XBB.1.5) vaccine in previously COVID-19 mRNA vaccinated and vaccine-naive participants: a 6-month follow-up

**DOI:** 10.1101/2025.04.09.25325550

**Authors:** Katia Alves, Karen Kotloff, R. Scott McClelland, E. Adrianne Hammershaimb, Alex Kouassi, Joyce S. Plested, Raj Kalkeri, Mingzhu Zhu, Shane Cloney-Clark, Zhaohui Cai, Katherine Smith, Muneer Kaba, Joy Nelson, Raburn M. Mallory, Fernando Noriega, the 2019nCoV-313 Study Investigators

**Author notes:** **Corresponding Author:** Katia Alves, MD, Novavax, Inc., 700 Quince Orchard Road, Gaithersburg, Maryland, 20878, USA.

## Abstract

**Background:** Seasonal (2023-2024) COVID-19 vaccine recommendations included updates against Omicron XBB.1.5. NVX-CoV2601 contains XBB.1.5 recombinant spike (rS) protein, Matrix-M™ adjuvant, and is based on authorized prototype vaccine (NVX-CoV2373) technology. Immunogenicity and safety outcomes 6 months after vaccination following a single dose of monovalent NVX-CoV2601 in previously vaccinated and vaccine-naive participants aged ≥18 years are reported here.

**Methods:** The phase 2/3 open-label, single-arm 2019nCoV-313 study consisted of two parts (part 1: participants with ≥3 prior mRNA vaccines; part 2: unvaccinated participants with a clinical history of COVID-19). Participants received a single dose of NVX-CoV2601. Primary endpoint analyses through day 28 were previously published. This final analysis assessed immunogenicity and safety through the end of the study (day 180). Immunogenicity data (e.g., neutralizing antibodies [nAbs] and anti-rS IgG antibodies) were summarized using geometric mean antibody levels and fold rise and seroresponse rate (SRR).

**Results:** The safety analysis set included 332 participants in part 1 and 338 participants in part 2. In previously vaccinated participants, nAb geometric mean titers (GMTs; 95% CIs) were 120.7 (101.5–143.6) at day 0, increased to 955.5 (814.0–1121.4) at day 28, and decreased to 454.8 (382.9–540.3) at day 180. SRR decreased from 64.3% at day 28 to 41.1% at day 180. Similar results were seen in vaccine-naive participants, with GMTs of 67.0 (56.6–79.3), 1296.7 (1082.6–1553.2), and 303.6 (258.5–356.4) at day 0, 28, and 180, respectively. SRR waned from 74.3% at day 28 to 45.0% at day 180. Anti-rS IgG responses similarly increased at day 28 and had moderate decreases at day 180 in both groups. No new safety signals were reported.

**Conclusions:** A single dose of NVX-CoV2601 showed robust, durable immunogenicity in both previously vaccinated and vaccine-naive adult participants. These data support the use of NVX-CoV2601 in both populations.

**Trial registration:** NCT05975060

**Highlights:**

- Single-dose XBB.1.5–based vaccine, NVX-CoV2601, was well-tolerated
- NVX-CoV2601 elicited robust immunogenicity, regardless of prior vaccination status
- Immune responses waned over time but remained well above baseline

## Introduction

Prototype COVID-19 vaccines, including BNT162b2 (Pfizer-BioNTech)^1^, mRNA-1273 (Moderna)^2^, and NVX-CoV2373 (Matrix-M™–adjuvanted, recombinant spike [rS] protein vaccine; Nuvaxovid™, Novavax, Inc.)^3,4^ directed to the ancestral (Wuhan) strain provided high protective efficacy against symptomatic COVID-19 early in the pandemic. However, variants of severe acute respiratory syndrome coronavirus (SARS-CoV-2) have evolved rapidly.^5^ A series of vaccine updates were recommended by regulatory agencies and have been implemented to help combat these variants, including updates to vaccines against the Omicron XBB.1.5 subvariant for the 2023–2024 season.^6^ As part of this effort, the NVX-CoV2601 vaccine targeting the XBB.1.5 rS protein was produced using the same technology as that used previously to produce the ancestral prototype vaccine NVX-CoV2373. NVX-CoV2601 was authorized for use in 2023–2024 in Canada, the European Union, the United States (US), and by the World Health Organization.^7,8^

Vaccination subsequent to the primary series (with any of the authorized/approved vaccines) is estimated to have been received by 32% of the global population, while approximately 20% of the population remains unvaccinated but has been exposed to SARS-CoV-2.^9^ The immunogenicity and safety of a single dose of NVX-CoV2601 was investigated in a 2-part, phase 2/3, open-label study (NCT05975060)^10^ that assessed outcomes in two relevant adult populations; part 1 included participants who had previously received ≥3 doses of an mRNA-based COVID-19 vaccine; part 2 included vaccine-naive (i.e., unvaccinated) participants who had evidence of prior SARS-CoV-2 infection, as determined by the presence of anti-nucleocapsid [anti-N] antibodies).

As previously reported,^11,12^ an interim, 28-day analysis in each of the aforementioned study populations met their respective primary endpoints with robust immunogenicity responses to NVX-CoV2601. Exploratory analyses determined that NVX-CoV2601 elicited immune responses to variants circulating at the time of the report. This report describes final immunogenicity and safety outcomes, 6 months after vaccination with NVX-CoV2601 in participants who had previously received ≥3 doses of an mRNA-based COVID-19 vaccine and in vaccine-naive participants who had evidence of prior SARS-CoV-2 infection.

## Methods

### Study design and participants

Parts 1 and 2 of the 2019nCoV-313 (NCT05975060) phase 2/3, open-label, single-arm study were conducted across 30 US sites. Part 1 enrolled medically stable, non-pregnant individuals aged ≥18 years who had previously received ≥3 doses of mRNA-1273 or BNT162b2 monovalent or bivalent vaccines, with the last dose administered ≥90 days prior to study vaccination.^12^ Part 2 enrolled medically stable, non-pregnant individuals aged ≥18 years who never received a SARS-CoV-2 vaccine and who reported an episode of COVID-19–like disease during the previous year^11^. All participants provided written informed consent.

The study protocol was approved by Advarra, Inc. (Columbia, MD, USA), and the study was conducted according to the principles of the International Conference on Harmonization Good Clinical Practice guidelines as well as all applicable national, state, and local laws and regulations.

### Procedures

Participants in both parts of the study received a single 0.5-mL intramuscular injection of trial vaccine (NVX-CoV2601) on day 0 and remained on study for immunogenicity and safety data collection through day 180, with scheduled visits on days 28, 90 (phone visit), and 180. NVX-CoV2601 contains 5 μg rS protein from XBB.1.5 and 50 μg Matrix-M adjuvant. Study vaccine was manufactured by the Serum Institute of India, Pvt., Ltd. (Pune, Maharashtra, India) in collaboration with Novavax, Inc. Participants in parts 1 and 2 of the study received doses of NVX-CoV2601 from the same vaccine lot and remained in the clinic or under observation for ≥15 min post vaccination for monitoring.

The primary endpoint analyses through day 28 (interim analysis) were previously published.^11,12^ This report includes data collected through the end of the study (day 180). Safety parameters evaluated through the end of the study included serious TEAEs (SAEs), treatment-related medically attended adverse events (MAAEs), and adverse events of special interest (AESIs). MAAEs were defined as any TEAE occurring after study vaccination leading to an unscheduled visit to a healthcare practitioner. AESIs included potential immune-mediated medical conditions, myocarditis, pericarditis, complications specific to COVID-19. TEAEs were categorized as SAEs if they were associated with death, hospitalization, persistent or significant incapacity or substantial disruption to normal life, congenital anomaly, or other serious, important medical events. Solicited local and systemic AEs were collected through day 7; treatment-related unsolicited treatment emergent adverse events (TEAEs) were collected through day 28. These data were previously published as part of the 28-day interim analyses.^11,12^

Participants were tested for SARS-CoV-2 infection at enrollment (day 0) by polymerase chain reaction of a nasal swab sample. Blood samples for immunogenicity assessments were collected on days 0, 28, and 180, and were evaluated centrally (Novavax Clinical Immunology, Gaithersburg, MD, USA). Serum was analyzed using a validated pseudovirus neutralization assay with an inhibitory dilution of 50% (ID_50_),^13^ anti-rS immunoglobulin G (IgG) enzyme-linked immunosorbent assay (ELISA),^14^ and anti-N serology.^15^

### Statistical analysis

Immunogenicity data were summarized using geometric mean titers (GMTs) and ELISA units (GMEUs) for neutralizing antibodies (nAbs) and anti-rS IgG antibodies, respectively, and geometric mean fold rise (GMFR) and seroresponse rate (SRR). GMTs and GMEUs were calculated as the antilog of the mean of the -log-transformed value at day 28 or day 180 to generate a normal distribution. SRR was defined as the percentage of participants with baseline result equal or greater than the lower limit of quantification (LLOQ) who achieved a ≥4-fold increase in antibody response from baseline, or for those with baseline <LLOQ, a post baseline response of ≥4-fold LLOQ. An analysis of covariance (ANCOVA) with vaccine group as fixed effect and baseline value (day 0) as covariate was performed to estimate adjusted GMTs/GMEUs and corresponding ratios (GMTR/GMEUR). The mean difference between vaccine groups and the corresponding CI limits were exponentiated to obtain the ratios and corresponding 95% CIs.

Descriptive methods were used to assess vaccine safety using a data set that included all participants who received the study vaccine. SAS® software (version 9.4 or higher; SAS Institute Inc., Cary, NC, USA) was used for all statistical analyses.

## Results

### Participants

The full analysis sets consisted of 332 participants enrolled into part 1 and 338 participants enrolled into part 2. The per-protocol analysis sets for the end-of-study/day 180 analysis included 311/332 (93.7%) participants in part 1 and 309/338 (91.4%) participants in part 2 (**Figure S1**). Some differences in participant demographics between the two parts of this study were noted (**Table 1**). In the per-protocol analysis set, part 1 vs part 2 participants were generally older (mean age, 52.1 vs 40.1 years), and a larger proportion were White (74.9% vs 49.5%). There was a larger proportion of female versus male participants in both parts of the study. Anti-N antibody was detected in all part 2 participants, per protocol, as well as 216/311 (69.5%) participants in part 1. Per study criteria, all participants in part 2 of the study were seropositive for SARS-CoV-2. Baseline demographics and participant characteristics were similar between the per-protocol and safety analysis sets (**Table S1**).

**Table 1.** Participant baseline demographics and characteristics in the per-protocol analysis set.

|  | **Part 1:**<br>**Previously vaccinated**<br>**(N=311)** | **Part 2:**<br>**Vaccine-naïve**<br>**(N=306)<sup>a</sup>** |
| --- | --- | --- |
| **Age, years** |  |  |
| Mean (SD) | 52.1 (16.1) | 40.1 (12.91) |
| Median (range) | 53 (18-89) | 38 (18-75) |
| **Age, category, n (%)** |  |  |
| 18–54 years | 164 (52.7) | 263 (85.9) |
| ≥55 years | 147 (47.3) | 43 (14.1) |
| **Sex, n (%)** |  |  |
| Female | 193 (62.1) | 177 (57.8) |
| Male | 118 (37.9) | 129 (42.2) |
| **Race, n (%)** |  |  |
| White | 233 (74.9) | 152 (49.7) |
| Black or African American | 47 (15.1) | 132 (43.1) |
| Asian | 12 (3.9) | 2 (0.7) |
| Native American or Alaska Native | 6 (1.9) | 6 (2.0) |
| Native Hawaiian or other Pacific Islander | 2 (0.6) | 1 (0.3) |
| Multiple | 3 (1.0) | 5 (1.6) |
| Other | 1 (0.3) | 2 (0.7) |
| Unknown | 1 (0.3) | 0 |
| Not reported | 6 (1.9) | 6 (2.0) |
| **Ethnicity, n (%)** |  |  |
| Not Hispanic or Latino | 246 (79.1) | 224 (73.2) |
| Hispanic or Latino | 62 (19.9) | 80 (26.1) |
| Not reported | 3 (1.0) | 2 (0.7) |
| **Previous COVID-19 vaccine doses<sup>b</sup>, n (%)** |  |  |
| 3 doses | 129 (41.5) | — |
| 4 doses | 109 (35.0) | — |
| 5 doses | 65 (20.9) | — |
| 6 doses | 8 (2.6) | — |
| **PCR, n (%)** |  |  |
| Negative | 311 (100)<sup>c</sup> | 306 (100)<sup>d</sup> |
| Positive | 0 | 0 |
| **Anti-N status, n (%)** |  |  |
| Positive | 216 (69.5) | 306 (100) |
| Negative | 95 (30.5) | 0 |

COVID-19 = coronavirus disease 2019; N=nucleocapsid; NR=not reported; PCR=polymerase chain reaction; SD=standard deviation.
<sup>a</sup>Day 28 Interim Analysis.
<sup>b</sup>Includes homologous and heterologous doses of either mRNA vaccine.
<sup>c</sup>Two participants with missing PCR status at baseline were imputed as negative.
<sup>d</sup>Five participants with missing PCR status at baseline were imputed as negative in part 2.

### Immunogenicity

In part 1, previously vaccinated participants exhibited an increase in nAbs against XBB.1.5 from day 0 (GMT: 120.7 [95% CI: 101.5–143.6]) to day 28 (955.5 [814.0–1121.4]). A decline was observed on day 180 (454.8 [382.9–540.3]), but GMTs remained substantially above baseline levels (**Figure 1**). GMFR from day 0 was 7.9 (95% CI: 6.8–9.2) at day 28 and 3.8 (3.2–4.4) at day 180. The SRR was 64.3% at day 28 and 41.1% at day 180. This pattern was consistent when analyzed by age group (18 to 54 years vs ≥55 years) and by the specific formulation of mRNA vaccine previously received (BNT162b2 vs mRNA-1273).

**Figure 1.**
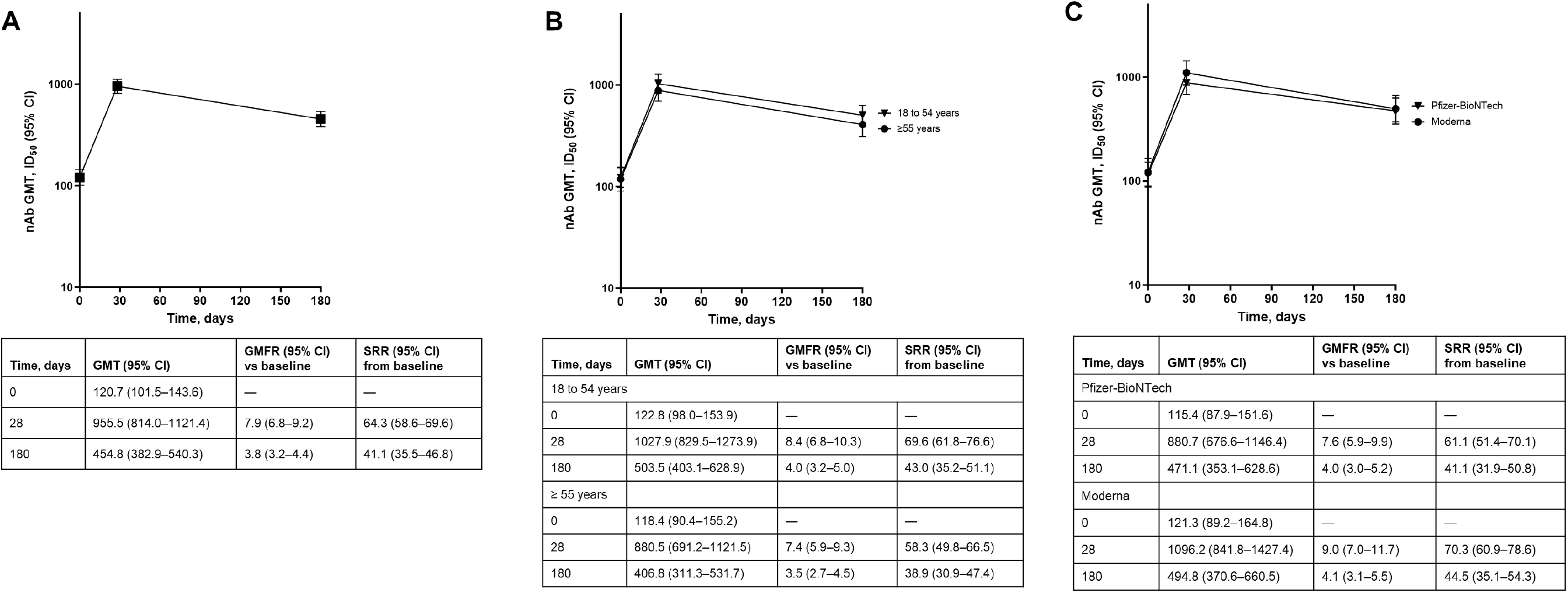
GMT (95% CI) of nAb responses to XBB.1.5 in previously vaccinated participants who received NVX-CoV2601 in 2019nCoV-313 part 1 (per-protocol analysis set) for (A) all participants, (B) analyzed by age group, and (C) analyzed by prior mRNA vaccine. GMT=geometric mean titer; GMFR=geometric mean fold rise; ID_50_=inhibitory dilution of 50%; mRNA=messenger ribonucleic acid; nAb=neutralizing antibody; SRR=seroresponse rate.

In part 2, vaccine-naive participants exhibited a similar pattern of increased nAb responses from day 0 (GMT: 67.0 [95% CI: 56.6–79.3]) to day 28 (1296.7 [1082.6–1553.2), then decreased by day 180 (303.6 [258.5–356.4]) but remained above baseline levels (**Figure 2**). GMFR from day 0 was 19.3 (95% CI: 15.7–23.7) at day 28 and 4.7 (3.8–5.7) at day 180. The SRR similarly decreased from 74.3% at day 28 to 45.0% at day 180. This pattern was consistent when analyzed by age group (18 to 54 years vs ≥55 years).

**Figure 2.**
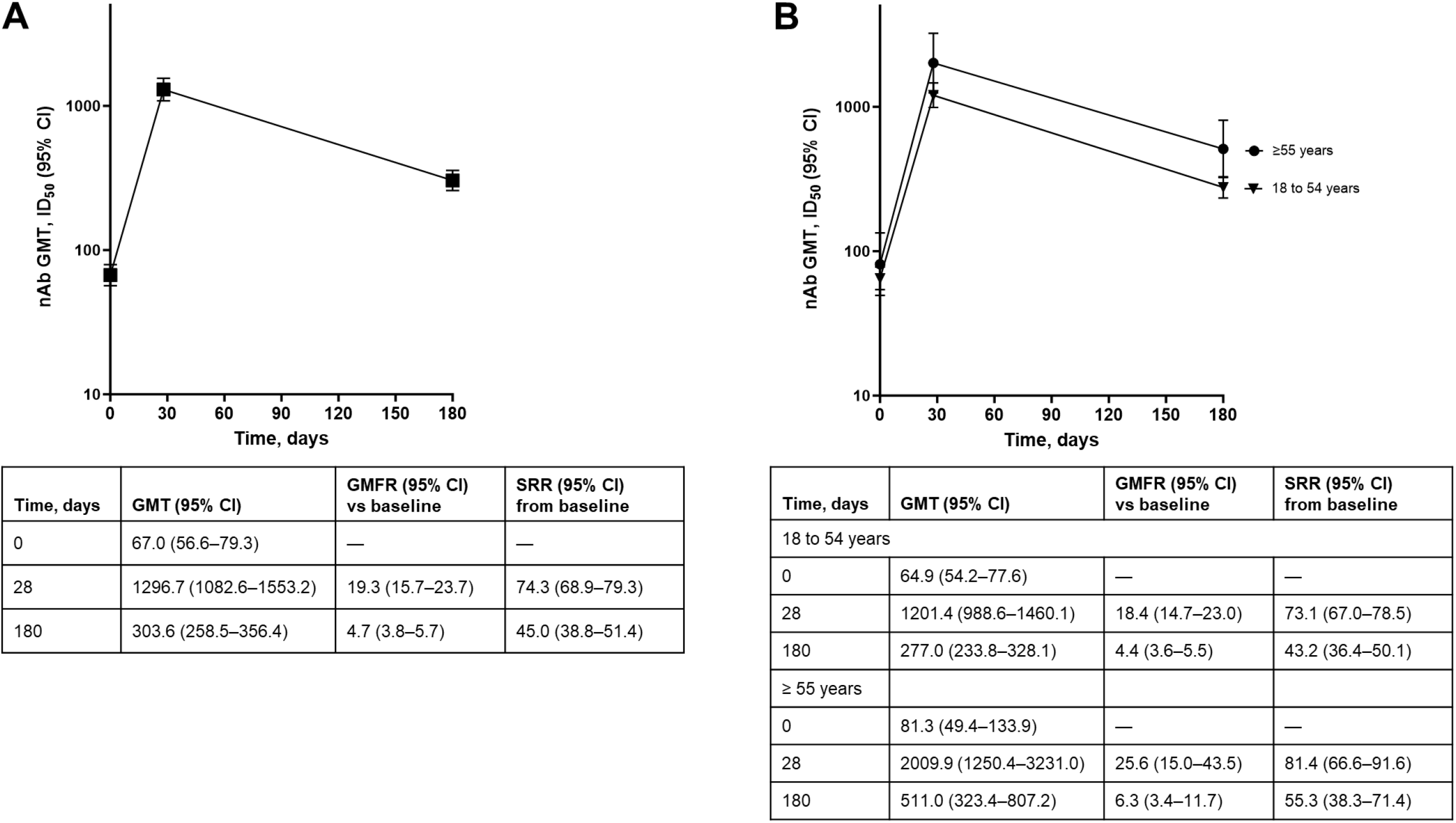
GMT (95% CI) of nAb responses to XBB.1.5 in vaccine-naive participants who received NVX-CoV2601 in 2019nCoV-313 part 2 (per-protocol analysis aet) for (A) all participants and (B) analyzed by age group. CI=confidence interval; GMT=geometric mean titer; GMFR=geometric mean fold rise; ID_50_=inhibitory dilution of 50%; nAb=neutralizing antibody; SRR=seroresponse rate.

Anti-rS IgG responses against XBB.1.5 followed a similar pattern of robust responses at day 28 that were lower at day 180, for both previously vaccinated (**Figure S2**) and vaccine-naive participants (**Figure S3**).

### Safety

Overall, unsolicited TEAEs were reported in 30/332 (9.0%) and 18/338 (5.3%) previously vaccinated and vaccine-naive participants, respectively (**Table 2**), with few participants reporting treatment-related unsolicited TEAEs (1.2% and 0.3%, respectively). MAAEs were reported in 14/332 (4.2%) and 8/338 (2.4%) previously vaccinated and vaccine-naive participants, respectively (**Table 2**). One MAAE of grade 2 hypertension in the previously vaccinated group was considered treatment-related (**Table S2**). SAEs and severe TEAEs were reported in <2% of participants in both the previously vaccinated and vaccine-naive groups, none of which were considered treatment-related (**Table 2**). Newly reported SAEs since the 28-day interim analysis^11,12^ included two in previously vaccinated participants (migraine and ureterolithiasis) and four in vaccine-naive participants (partial large bowel obstruction, acute myocardial infarction, myocardial infarction, and fentanyl overdose); each were considered by the investigators to be unrelated to treatment.

**Table 2.** Summary of unsolicited AEs in participants vaccinated with NVX-CoV2601 (XBB.1.5) (safety analysis set)

|  | **Part 1:**<br>**Previously vaccinated**<br>**(N=332)** | **Part 2:**<br>**Vaccine-naïve**<br>**(N=338)** |
| --- | --- | --- |
| Unsolicited TEAEs, n (%)<sup>a</sup> |  |  |
| Any<sup>a</sup> | 30 (9.0) | 18 (5.3) |
| Treatment-related non-serious<sup>a</sup> | 4 (1.2) | 1 (0.3) |
| Severe TEAEs, n (%)<sup>a</sup> | 2 (0.6) | 1 (0.3) |
| Treatment-related<sup>a</sup> | 0 | 0 |
| SAE, n (%) | 4 (1.2)<sup>a</sup> | 5 (1.5)<sup>b</sup> |
| Treatment-related | 0<sup>a</sup> | 0<sup>b</sup> |
| MAAE, n (%)<sup>a</sup> | 14 (4.2) | 8 (2.4) |
| Treatment-related<sup>b</sup> | 0 | 0 |
| AESIs (PIMMCs), n (%)<sup>b</sup> | 0 | 0 |
| AESIs: Relevant to COVID-19<sup>b</sup> |  | 0 |
| Any Myocarditis/Pericarditis<sup>b</sup> |  | 0 |
| TEAEs leading to study discontinuation, n (%)<sup>b</sup> | 0 | 1 (0.3) |
| Death, n (%) | 0 | 1 (0.3) |

AE=adverse event; AESI=AE of special interest; MAAE=medically attended AE; PIMMC=potential immune-mediated medical condition; SAE, serious AE; TEAE=treatment emergent AE.
<sup>a</sup>Through 28 days post vaccination.
<sup>b</sup>Through end of study.

No participant reported a TEAE of myocarditis/pericarditis (**Table 2**). There was one death reported in part 2 of the study, which occurred in a vaccine-naive participant who died of a fentanyl overdose; this death was assessed by the investigator and sponsor as unrelated to the study vaccine. Other than this one death reported in part 2 of the study, no participants discontinued the study due to a TEAE.

## Discussion

In this end-of-study analysis, a single dose of the monovalent XBB.1.5 subvariant vaccine, NVX-CoV2601, induced a robust nAb response that maintained levels at day 180 that were well above baseline; although decreased over time in both previously vaccinated and vaccine-naive participants. Additionally, no new safety signals since the 28-day interim analysis^11,12^ were identified in the analysis of safety and reactogenicity.

Both groups of participants demonstrated a decrease in a nAb response from day 28 until the end of the study period, regardless of age (both groups) and number/type of prior mRNA vaccination (previously vaccinated group). In previously vaccinated participants, the nAb response decreased by approximately 50% from day 28 to day 180; in vaccine-naive participants, the nAb response decreased by approximately 76% in the same time frame. There was a more robust initial nAb response observed from day 0 to day 28 for the vaccine-naive versus previously vaccinated groups (9.0-fold vs 3.5-fold); therefore, allowing for an opportunity to have a greater drop in nAb response from day 28 to day 180 in these respective groups. However, the gradual decline in immune response over time is a well-documented characteristic of certain subsets of vaccines.^16^ COVID-19 vaccines, similar to influenza vaccines, provide a shorter duration of protection (<5 years) than other vaccines that provide a moderate (5–20 years; e.g., human papillomavirus and tetanus vaccines) or long (>20 years; e.g., measles and rubella vaccines) duration of protection.^16^ Evidence thus far suggests that neither SARS-CoV-2 infection nor COVID-19 vaccines provide lasting protection; however, this can largely be attributed to the rapid mutation rate and immune evasive nature of SARS-CoV-2,^17-19^ as well as the low replication fidelity of the RNA-dependent RNA polymerase^20^ involved in proliferation of SARS-CoV-2. Most studies, including systematic reviews, have demonstrated significantly waning protection within 3–6 months, especially against Omicron variants.^16^ One review evaluating 78 studies on vaccine efficacy before the advent of the Omicron variant concluded that vaccine efficacy against symptomatic disease waned by 20% to 30% 6 months after a primary series.^21^ In fact, in the PREVENT-19 trial, vaccine efficacy against the Delta (i.e., B.1.617.2) strain was 88%, 82%, and 77% at 40, 120, and 180 days, respectively, with evidence of waning (*P* < 0.01) after two doses of NVX-CoV2373.^22^ Another review based on 40 studies identified a faster waning of vaccine efficacy against the Omicron than the Delta variant.^18^ Importantly, the nAb response has been assessed as a correlate of protection against COVID-19.^23^ Fong et al. showed that a 50% pseudovirus nAb titer measured 2 weeks after the second dose of NVX-CoV2373 was inversely associated with COVID-19 risk and vaccine efficacy, with vaccine efficacy estimates of 75.7%, 81.7%, and 96.8% in vaccine recipients with titers of 50, 100, and 7230 international units/mL, respectively.^23^ Although this study by Fong et al. predated the emergence of Delta and Omicron variants of concern, the nAb response may be considered a correlate of protection for the NVX-CoV2601 vaccine, especially as the response displayed by both previously vaccinated and vaccine-naive participants in the current study was maintained substantially above baseline levels 6 months after a single dose.

The 2-part design of this study allowed investigation of outcomes in previously vaccinated and unvaccinated representative populations. A single dose of NVX-CoV2601 led to a similar nAb response in both study populations. Fold increases from day 0 to day 28 and from day 28 to day 180 were more pronounced in vaccine-naive versus previously vaccinated participant group; however, this difference may be due to lower baseline antibody levels in the former population. Outcomes in previously vaccinated participants indicate that updating the COVID-19 vaccine to include XBB.1.5 produces robust immune responses through 180 days post vaccination. Furthermore, the robust responses observed in vaccine-naive participants demonstrate that a single dose of NVX-CoV2601 was sufficient to augment immunogenicity derived from natural infection. This finding is in line with other studies that have shown a single vaccine dose received by an unvaccinated individual post SARS-CoV-2 infection can induce a comparable, and possibly greater, immune response, compared with individuals who received two vaccine doses.^24,25^ As the majority of the population (>96% aged ≥16 years) can be presumed to have SARS-CoV-2 antibodies via either vaccination and/or exposure^26^, results of the current study support the use of NVX-CoV2601 as a single dose in both vaccinated and unvaccinated individuals.

The results from this end-of-study analysis describes durability of the nAb response and adds to the findings from the 28-day interim data,^11,12^ as well as to the findings from other studies investigating the immunogenicity and effectiveness of updated monovalent vaccines directed against XBB.1.5.^27-32^ In September 2023, the US FDA authorized the use of COVID-19 mRNA-based vaccines directed to XBB.1.5 (in individuals aged ≥6 months), as well as NVX-CoV2601 (in individuals aged ≥12 years); NVX-CoV2601 received EMA authorization shortly thereafter, in October 2023 (US FDA press release, 2023; EMA, 2023, 2024). Previous investigations of updated monovalent mRNA vaccines showed notable nAb responses against XBB.1.5, similar to those described here, albeit with a smaller population than that analyzed in this study.^27,28,32^

Safety of the NVX-CoV2601 vaccine in this study was consistent with that described previously for the ancestral strain vaccine NVX-CoV2373.^3,4,33,34^ Most of the unsolicited AEs occurred during the first 28 days post vaccination.^11,12^ More unsolicited AEs were reported in participants from part 1 than in part 2, which may be related to the higher mean participant age in part 1 compared to part 2, which tend to occur more frequently in an aging population.^35^ Six new SAEs were reported since the 28-day interim analysis,^11,12^ none of which were considered to be related to treatment; one fatal fentanyl overdose occurred in a vaccine-naive participant.

This analysis is potentially limited by some demographic differences between the previously vaccinated and the vaccine-naive participants, including differences in age, sex, and race, which likely reflect differences in the vaccinated and unvaccinated populations in the US.^36^ Despite these differences, immunogenicity and safety outcomes were consistent across the vaccine groups. Additionally, a large proportion (>65%) of the previously vaccinated participants had prior SARS-CoV-2 infection; thus, possible effects of hybrid immunity cannot be ruled out in the part 1 population.

In summary, the 2019nCoV-313 study demonstrated robust immunogenicity through 180 days post vaccination with a single dose of NVX-CoV2601 in both previously vaccinated and vaccine-naive participants, and no new safety signals were observed. Together, these long-term data support the use of the Novavax vaccine platform in these two populations.

## Data Availability

Study information is available online at https://www.clinicaltrials.gov/study/NCT05975060. Requests submitted to the corresponding author will be considered upon publication of this article and de-identified participant data, related to results reported here, would be provided. The study protocol is available in supplemental material.

## Acknowledgments

Medical writing support for the development of this manuscript, under the direction of the authors, was provided by Kelly M. Cameron, PhD, CMPP, Liana E. Merrill, PhD, and Ebenezer M. Awuah-Yeboah, BS of Ashfield MedComms (US), an Inizio company, and was funded by Novavax, Inc. The 2019nCoV-313 study was funded by Novavax, Inc., who provided the vaccine, collaborated with the investigators on protocol design, data analysis and interpretation, and preparation of this report.

## Author Contributions

KA, KK, RMM, and FN were involved in the study design and conducted the data analysis and interpretation. KK, RSM, and EAH were involved in data collection. JN provided project management. AK performed the statistical analyses. KA, RSM, AK, and FN have directly accessed and verified the data in this manuscript. KA, KK, RSM, AK, JSP, RK, MZ, SC-C, ZC, KS, MK, JN, EAH, RMM, and FN were involved in data interpretation and reviewed, commented on, and approved this manuscript prior to submission for publication. The authors accept accountabilities for all aspects of the work, ensuring questions related to accuracy or integrity are investigated and resolved. All authors had full access to all the data in the study and had final responsibility for the decision to submit for publication.

## References

1. Polack FP, Thomas SJ, Kitchin N, et al. Safety and Efficacy of the BNT162b2 mRNA Covid-19 Vaccine. N Engl J Med 2020; 383(27): 2603–15.

2. Baden LR, El Sahly HM, Essink B, et al. Efficacy and Safety of the mRNA-1273 SARS-CoV-2 Vaccine. N Engl J Med 2021; 384(5): 403–16.

3. Dunkle LM, Kotloff KL, Gay CL, et al. Efficacy and Safety of NVX-CoV2373 in Adults in the United States and Mexico. N Engl J Med 2022; 386(6): 531–43.

4. Heath PT, Galiza EP, Baxter DN, et al. Safety and Efficacy of NVX-CoV2373 Covid-19 Vaccine. N Engl J Med 2021; 385(13): 1172–83.

5. Ao D, He X, Hong W, Wei X. The rapid rise of SARS-CoV-2 Omicron subvariants with immune evasion properties: XBB.1.5 and BQ.1.1 subvariants. MedComm (2020) 2023; 4(2): e239.

6. World Health Organization. Statement on the antigen composition of COVID-19 vaccines. 18 May 2023. https://www.who.int/news/item/18-05-2023-statement-on-the-antigen-composition-of-covid-19-vaccines (accessed 27 March 2024).

7. World Health Organization. COVID-19 Vaccines with WHO Emergency Use Listing. https://extranet.who.int/prequal/vaccines/covid-19-vaccines-who-emergency-use-listing (accessed 27 March 2024).

8. Novavax. Novavax’s Updated COVID-19 Vaccine Now Authorized in Canada. 5 December 2023. https://ir.novavax.com/press-releases/2023-12-05-Novavaxs-Updated-COVID-19-Vaccine-Now-Authorized-in-Canada (accessed 27 March 2024).

9. World Health Organization. COVID-19 vaccination, United States of America data. 2023. https://data.who.int/dashboards/covid19/vaccines?m49=840 (accessed January 13 2025).

10. NIH National Library of Medicine. Clinicaltrials.gov. NCT05975060. May 30, 2024. https://clinicaltrials.gov/study/NCT05975060 (accessed November 13, 2024.

11. Alves K, Kouassi A, Plested JS, et al. Immunogenicity and safety of a monovalent Omicron XBB.1.5 SARS-CoV-2 recombinant spike protein vaccine in previously unvaccinated, SARS-CoV-2 seropositive participants: primary day-28 analysis of a phase 2/3 open-label study. Vaccine. 2025;55:127046.

12. Alves K, Kotloff K, McClelland RS, et al. Immunogenicity and safety of a monovalent omicron XBB.1.5 SARS-CoV-2 recombinant spike protein vaccine as a heterologous booster dose in US adults: interim analysis of a single-arm phase 2/3 study. Lancet Infect Dis 2025.

13. Cai Z, Kalkeri R, Wang M, et al. Validation of a Pseudovirus Neutralization Assay for Severe Acute Respiratory Syndrome Coronavirus 2: A High-Throughput Method for the Evaluation of Vaccine Immunogenicity. Microorganisms 2024; 12(6).

14. Zhu M, Cloney-Clark S, Feng SL, et al. A Severe Acute Respiratory Syndrome Coronavirus 2 Anti-Spike Immunoglobulin G Assay: A Robust Method for Evaluation of Vaccine Immunogenicity Using an Established Correlate of Protection. Microorganisms 2023; 11(7).

15. Madhi SA, Moodley D, Hanley S, et al. Immunogenicity and safety of a SARS-CoV-2 recombinant spike protein nanoparticle vaccine in people living with and without HIV-1 infection: a randomised, controlled, phase 2A/2B trial. Lancet HIV 2022; 9(5): e309–e22.

16. Vashishtha VM, Kumar P. The durability of vaccine-induced protection: an overview. Expert Rev Vaccines 2024; 23(1): 389–408.

17. Carabelli AM, Peacock TP, Thorne LG, et al. SARS-CoV-2 variant biology: immune escape, transmission and fitness. Nat Rev Microbiol 2023; 21(3): 162–77.

18. Menegale F, Manica M, Zardini A, et al. Evaluation of Waning of SARS-CoV-2 Vaccine-Induced Immunity: A Systematic Review and Meta-analysis. JAMA Netw Open 2023; 6(5): e2310650.

19. Reuschl AK, Thorne LG, Whelan MVX, et al. Evolution of enhanced innate immune suppression by SARS-CoV-2 Omicron subvariants. Nat Microbiol 2024; 9(2): 451–63.

20. Yin X, Popa H, Stapon A, Bouda E, Garcia-Diaz M. Fidelity of Ribonucleotide Incorporation by the SARS-CoV-2 Replication Complex. J Mol Biol 2023; 435(5): 167973.

21. Feikin DR, Higdon MM, Abu-Raddad LJ, et al. Duration of effectiveness of vaccines against SARS-CoV-2 infection and COVID-19 disease: results of a systematic review and meta-regression. Lancet 2022; 399(10328): 924–44.

22. Follmann D, Mateja A, Fay MP, et al. Durability of Protection Against COVID-19 Through the Delta Surge for the NVX-CoV2373 Vaccine. Clinical infectious diseases : an official publication of the Infectious Diseases Society of America 2024; 79(1): 78–85.

23. Fong Y, Huang Y, Benkeser D, et al. Immune correlates analysis of the PREVENT-19 COVID-19 vaccine efficacy clinical trial. Nat Commun 2023; 14(1): 331.

24. Krammer F, Srivastava K, Alshammary H, et al. Antibody Responses in Seropositive Persons after a Single Dose of SARS-CoV-2 mRNA Vaccine. N Engl J Med 2021; 384(14): 1372–4.

25. Zar HJ, MacGinty R, Workman L, et al. Natural and hybrid immunity following four COVID-19 waves: A prospective cohort study of mothers in South Africa. EClinicalMedicine 2022; 53: 101655.

26. Jones JM, Manrique IM, Stone MS, et al. Estimates of SARS-CoV-2 Seroprevalence and Incidence of Primary SARS-CoV-2 Infections Among Blood Donors, by COVID-19 Vaccination Status - United States, April 2021-September 2022. MMWR Morb Mortal Wkly Rep 2023; 72(22): 601–5.

27. Chalkias S, McGhee N, Whatley JL, et al. Interim Report of the Reactogenicity and Immunogenicity of Severe Acute Respiratory Syndrome Coronavirus 2 XBB-Containing Vaccines. J Infect Dis 2024; 230(2): e279–e86.

28. Gayed J, Bangad V, Xu X, et al. Immunogenicity of the Monovalent Omicron XBB.1.5-Adapted BNT162b2 COVID-19 Vaccine against XBB.1.5, BA.2.86, and JN.1 Sublineages: A Phase 2/3 Trial. Vaccines 2024; 12(7).

29. Link-Gelles R, Ciesla AA, Mak J, et al. Early Estimates of Updated 2023-2024 (Monovalent XBB.1.5) COVID-19 Vaccine Effectiveness Against Symptomatic SARS-CoV-2 Infection Attributable to Co-Circulating Omicron Variants Among Immunocompetent Adults - Increasing Community Access to Testing Program, United States, September 2023-January 2024. MMWR Morb Mortal Wkly Rep 2024; 73(4): 77–83.

30. Lustig Y, Barda N, Weiss-Ottolenghi Y, et al. Humoral response superiority of the monovalent XBB.1.5 over the bivalent BA.1 and BA.5 mRNA COVID-19 vaccines. Vaccine 2024; 42(22): 126010.

31. Marking U, Bladh O, Aguilera K, et al. Humoral immune responses to the monovalent XBB.1.5-adapted BNT162b2 mRNA booster in Sweden. Lancet Infect Dis 2024; 24(2): e80–e1.

32. Wang Q, Guo Y, Bowen A, et al. XBB.1.5 monovalent mRNA vaccine booster elicits robust neutralizing antibodies against XBB subvariants and JN.1. Cell Host Microbe 2024; 32(3): 315–21 e3.

33. Bennett C, Rivers EJ, Woo W, et al. Immunogenicity and safety of heterologous Omicron BA.1 and bivalent SARS-CoV-2 recombinant spike protein booster vaccines: a phase 3, randomized, clinical trial. J Infect Dis 2023.

34. Bennett C, Woo W, Bloch M, et al. Immunogenicity and safety of a bivalent (omicron BA.5 plus ancestral) SARS-CoV-2 recombinant spike protein vaccine as a heterologous booster dose: interim analysis of a phase 3, non-inferiority, randomised, clinical trial. Lancet Infect Dis 2024; 24(6): 581–93.

35. Smith K, Hegazy K, Cai MR, McKnight I, Rousculp MD, Alves K. Safety of the NVX-CoV2373 COVID-19 vaccine in randomized placebo-controlled clinical trials. Vaccine 2023; 41(26): 3930–6.

36. Pingali C, Meghani M, Razzaghi H, et al. COVID-19 Vaccination Coverage Among Insured Persons Aged >/=16 Years, by Race/Ethnicity and Other Selected Characteristics - Eight Integrated Health Care Organizations, United States, December 14, 2020-May 15, 2021. MMWR Morb Mortal Wkly Rep 2021; 70(28): 985–90.

