## Supplementary File for "Phase 2/3 open-label study on NVX-CoV-2601 (XBB.1.5) vaccine in previously COVID-19 mRNA vaccinated and vaccine-naive participants: a 6-month follow-up"

**SUPPLEMENTAL MATERIAL**

**Figure S1.** Participant Disposition in the 2019nCoV-313 study for parts 1 and 2.

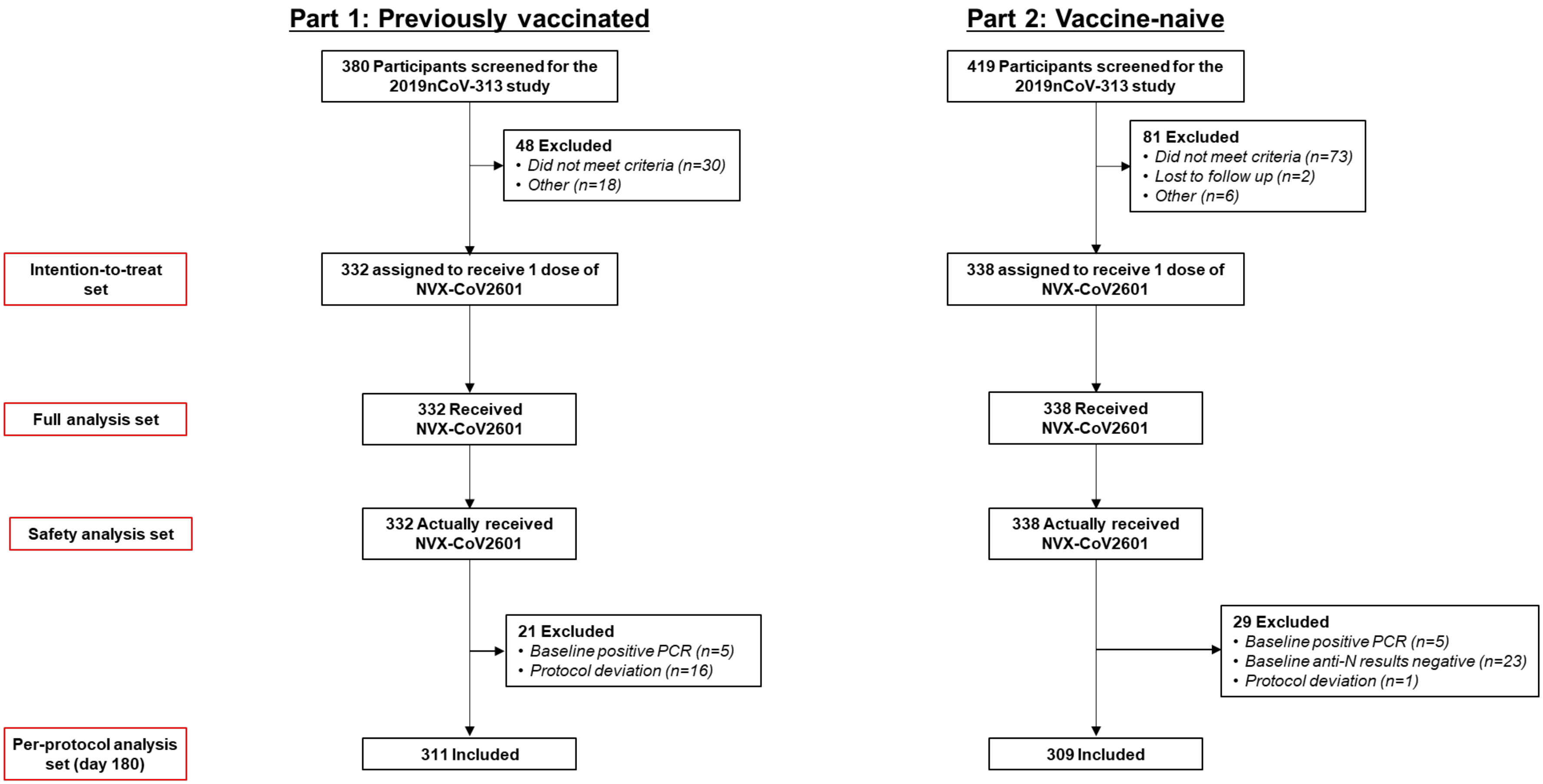


Participants may have been excluded for more than one reason.
PCR=polymerase chain reaction.

**Figure S2.** GMEU (95% CI) of anti-rS IgG responses to XBB.1.5 in previously vaccinated participants who received NVX-CoV2601 2019nCoV-313 part 1 (per protocol analysis set) for (A) all participants, (B) analyzed by age group, and (C) analyzed by prior mRNA vaccine. CI=confidence interval; ELISA=enzyme-linked immunosorbent assay; EU=ELISA units; GMEU=geometric mean ELISA units; GMFR=geometric mean fold rise; IgG=immunoglobulin G; mRNA=messenger ribonucleic acid.

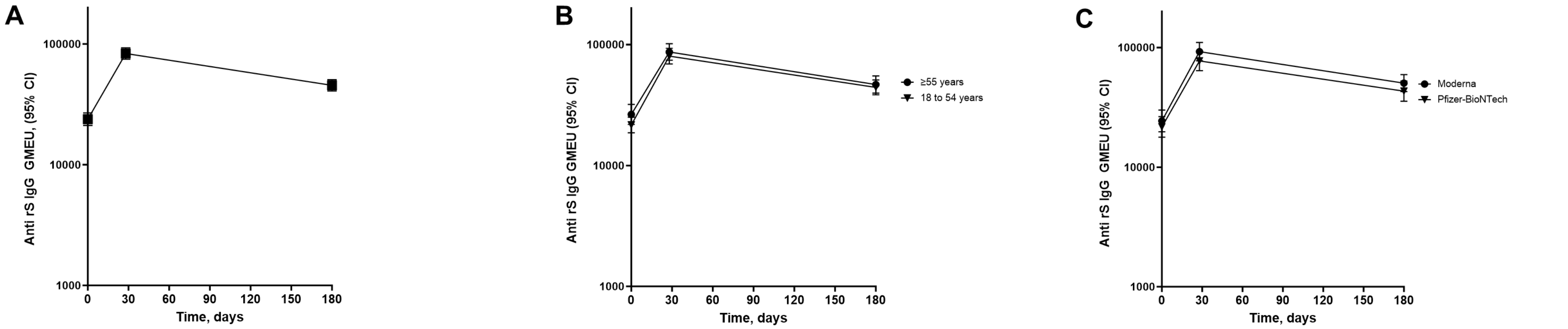


**A**

| Time, days | GMEU (95% CI) | GMFR (95% CI) vs baseline | SRR (95% CI) from baseline |
|---|---|---|---|
| 0 | 23771 (21110–26768) | — | — |
| 28 | 83206 (74747–92622) | 3.5 (3.2–3.9) | 39.7 (34.1–45.4) |
| 180 | 45388 (40799–50493) | 1.9 (1.7–2.1) | 17.5 (13.4–22.3) |

**B**

| Time, days | GMEU (95% CI) | GMFR (95% CI) vs baseline | SRR (95% CI) from baseline |
|---|---|---|---|
| 18 to 54 years | | | |
| 0 | 21611 (18656–25033) | — | — |
| 28 | 80163 (69114–92979) | 3.7 (3.2–4.2) | 44.1 (36.3–52.1) |
| 180 | 44242 (38419–50948) | 2.0 (1.8–2.3) | 17.7 (12.1–24.6) |
| ≥ 55 years | | | |
| 0 | 26425 (21841–31971) | — | — |
| 28 | 86744 (74160–101464) | 3.3 (2.8–3.9) | 34.7 (27.0–43.1) |
| 180 | 46680 (39663–54938) | 1.8 (1.5–2.1) | 17.4 (11.6–24.6) |

**C**

| Time, days | GMEU (95% CI) | GMFR (95% CI) vs baseline | SRR (95% CI) from baseline |
|---|---|---|---|
| Pfizer-BioNTech | | | |
| 0 | 21677 (17793–26409) | — | — |
| 28 | 77190 (64065–93003) | 3.6 (3.0–4.2) | 42.5 (33.2–52.1) |
| 180 | 43167 (35570–52388) | 1.9 (1.6–2.3) | 17.9 (11.3–26.2) |
| Moderna | | | |
| 0 | 24337 (19767–29965) | — | — |
| 28 | 92347 (77416–110158) | 3.8 (3.1–4.7) | 41.4 (32.2–51.2) |
| 180 | 50382 (42727–59408) | 2.1 (1.7–2.5) | 21.8 (14.5–30.7) |

**Figure S3.** GMEU (95% CI) of anti-rS IgG responses to XBB.1.5 in vaccine-naive participants who received NVX-CoV2601 in 2019nCoV-313 part 2 (per-protocol analysis set) for (A) all participants and (B) analyzed by age group. CI=confidence interval; ELISA=enzyme-linked immunosorbent assay; EU=ELISA units; GMEU=geometric mean ELISA units; GMFR=geometric mean fold rise; IgG=immunoglobulin G.

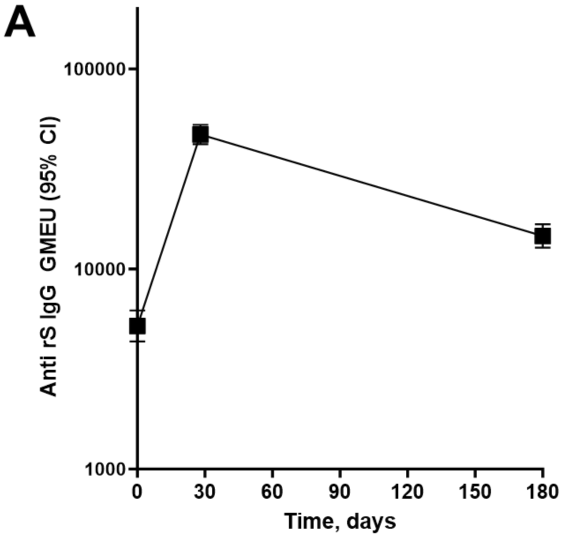


| Time, days | GMEU (95% CI) | GMFR (95% CI) vs baseline | SRR (95% CI) from baseline |
|---|---|---|---|
| 0 | 5188 (4342–6199) | — | — |
| 28 | 46962 (42047–52451) | 9.0 (7.6–10.7) | 68.4 (62.7–73.7) |
| 180 | 14607 (12772–16705) | 2.8 (2.4–3.4) | 38.2 (32.2–44.6) |

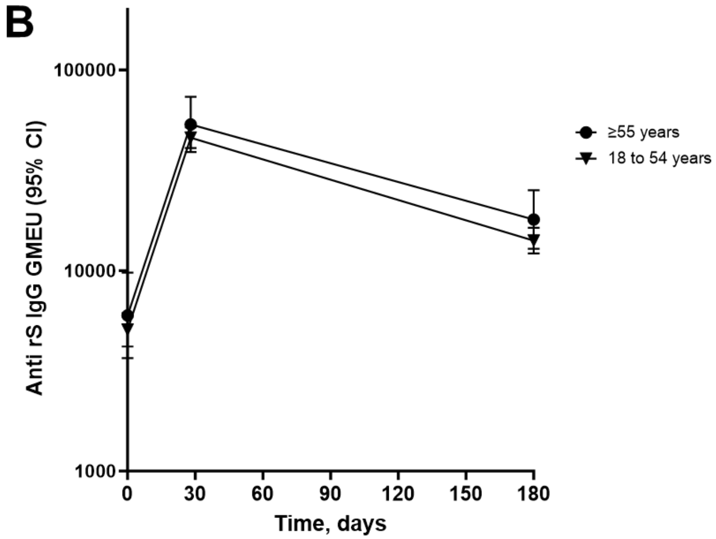


| Time, days | GMEU (95% CI) | GMFR (95% CI) vs baseline | SRR (95% CI) from baseline |
|---|---|---|---|
| 18 to 54 years | | | |
| 0 | 5067 (4181–6140) | — | — |
| 28 | 45910 (40788–51674) | 9.0 (7.4–10.9) | 67.8 (61.5–73.6) |
| 180 | 14087 (12163–16316) | 2.8 (2.3–3.4) | 37.6 (31.0–44.4) |
| ≥ 55 years | | | |
| 0 | 5968 (3656–9741) | — | — |
| 28 | 53487 (38977–73399) | 9.3 (6.4–13.6) | 72.1 (56.3–84.7) |
| 180 | 17947 (12824–25118) | 2.9 (1.9–4.5) | 42.1 (26.3–59.2) |

**Table S1.** Participant baseline demographics and characteristics in the safety analysis sets

| | Part 1: Previously vaccinated (N=332) | Part 2: Vaccine-naive (N=338) |
|---|---|---|
| **Age, years** | | |
| Mean (SD) | 52.0 (16.1) | 40.5 (13.11) |
| Median (range) | 53.0 (18-89) | 38.0 (18-75) |
| **Age, category, n (%)** | | |
| 18–54 years | 176 (53.0) | 284 (84.0) |
| ≥55 years | 156 (47.0) | 54 (16.0) |
| **Sex, n (%)** | | |
| Female | 208 (62.7) | 190 (56.2) |
| Male | 124 (37.3) | 148 (43.8) |
| **Race, n (%)** | | |
| White | 248 (74.7) | 167 (49.4) |
| Black or African American | 53 (16.0) | 147 (43.5) |
| Asian | 12 (3.6) | 2 (0.6) |
| Native American or Alaska Native | 6 (1.8) | 6 (1.8) |
| Native Hawaiian or other Pacific Islander | 2 (0.6) | 1 (0.3) |
| Multiple | 3 (0.9) | 5 (1.5) |
| Other | 1 (0.3) | 3 (0.9) |
| Unknown | 1 (0.3) | 0 |
| Not reported | 6 (1.8) | 7 (2.1) |
| **Ethnicity, n (%)** | | |
| Not Hispanic or Latino | 261 (78.6) | 249 (73.7) |
| Hispanic or Latino | 67 (20.2) | 87 (25.7) |
| Not reported | 4 (1.2) | 2 (0.6) |
| Unknown | 1 (0.3) | |
| **Previous COVID-19 vaccine doses[a], n (%)** | | |
| 3 doses | 135 (40.7) | — |
| 4 doses | 117 (35.2) | — |
| 5 doses | 70 (21.1) | — |
| 6 doses | 9 (2.7) | — |
| **PCR, n (%)** | | |
| Negative | 327 (98.5)[b] | 333 (98.5)[c] |
| Positive | 5 (1.5) | 5 (1.5) |
| **Anti-N status, n (%)** | | |
| Positive | 234 (70.5) | 315 (93.2) |
| Negative | 98 (29.5) | 23 (6.8) |

N=nucleocapsid; NR=not reported; PCR=polymerase chain reaction; SD=standard deviation.

[a]Includes homologous and heterologous doses of either mRNA vaccine.

[b]Two participants with missing PCR status at baseline were imputed as negative.

[c]Five participants with positive PCR results at baseline were excluded from the per-protocol analysis set.

**Table S2.** Unsolicited severe TEAEs, SAEs, MAAEs, and PIMMCs through the end of the study (safety analysis set)

| **System organ class***<br>Preferred term | **Part 1:<br>Previously vaccinated<br>(N=332)** | **Part 2:<br>Vaccine-naive<br>(N=338)** |
|---|---|---|
| **SAEs, n participants (%); n events** | | |
| Any SAE | 4 (1.2); 4 | 5 (1.5); 5 |
| Infections and infestations | 1 (0.3); 1 | 0 |
| Appendiceal abscess | 1 (0.3); 1 | 0 |
| Neoplasms, benign, malignant, and unspecified (including cysts and polyps) | 1 (0.3); 1 | 0 |
| Gastrointestinal stromal tumor | 1 (0.3); 1 | 0 |
| Nervous system disorders | 1 (0.3); 1 | 0 |
| Migraine | 1 (0.3); 1 | 0 |
| Renal and urinary disorders | 1 (0.3); 1 | 0 |
| Ureterolithiasis | 1 (0.3); 1 | 0 |
| Cardiac disorders | 0 | 2 (0.6); 2 |
| Acute myocardial infarction | 0 | 1 (0.3); 1 |
| Myocardial infarction | 0 | 1 (0.3); 1 |
| Gastrointestinal disorders | 0 | 2 (0.6); 2 |
| Large intestinal obstruction | 0 | 1 (0.3); 1 |
| Obstructive pancreatitis | 0 | 1 (0.3); 1 |
| Injury, poisoning and procedural complications | 0 | 1 (0.3); 1 |
| Overdose | 0 | 1 (0.3); 1 |
| **MAAEs, n participants (%); n events** | | |
| Any MAAE | 14 (4.2); 15 | 8 (2.4); 22 |
| Infections and infestations | 6 (1.8); 7 | 3 (0.9); 6 |
| Tooth abscess | 2 (0.6); 2 | 0 |
| Appendiceal abscess | 1 (0.3); 1 | 0 |
| COVID-19 | 1 (0.3); 1 | 0 |
| Gastroenteritis (*E. coli*) | 1 (0.3); 1 | 0 |
| Viral respiratory tract infection | 1 (0.3); 1 | 0 |
| Tonsillitis | 1 (0.3); 1 | 0 |
| Cellulitis | 0 | 1 (0.3); 1 |
| Otitis media | 0 | 1 (0.3); 1 |
| Urinary tract infection | 0 | 1 (0.3); 1 |
| Nervous system disorders | 2 (0.6); 2 | 0 |
| Migraine | 2 (0.6); 2 | 0 |
| Tension headache | 0 | 0 |
| Vascular disorders | 2 (0.6); 2 | 0 |
| Hypertension | 2 (0.6); 2 | 0 |
| Injury, poisoning, and procedural complications | 1 (0.3); 1 | 2 (0.6); 3 |
| Tooth fracture | 1 (0.3); 1 | 0 |
| Fractured coccyx | 0 | 1 (0.3); 1 |
| Post-traumatic pain | 0 | 1 (0.3); 1 |
| Road traffic accident | 0 | 1 (0.3); 1 |
| Investigations | 1 (0.3); 1 | 0 |
| Blood estrogen decreased | 1 (0.3); 1 | 0 |
| Neoplasms, benign and unspecified (incl. cysts and polyps) | 1 (0.3); 1 | 0 |
| Gastrointestinal stromal tumor | 1 (0.3); 1 | 0 |

| | | |
|---|---|---|
| Respiratory, thoracic and mediastinal disorders | 1 (0.3); 1 | 1 (0.3); 1 |
| Cough | 1 (0.3); 1 | 1 (0.3); 1 |
| Gastrointestinal disorders | 0 | 2 (0.6); 2 |
| Abdominal pain | 0 | 1 (0.3); 1 |
| Obstructive pancreatitis | 0 | 1 (0.3); 1 |
| Immune system disorders | 0 | 1 (0.3); 1 |
| Hypersensitivity | 0 | 1 (0.3); 1 |
| Psychiatric disorders | 0 | 1 (0.3); 1 |
| Anxiety | 0 | 1 (0.3); 1 |
| **PIMMCs, n participants (%), n events** | | |
| Any PIMMC | 0 | 0 |

*MedDRA version 25.0.

MAAE=medically attended adverse event; mRNA=messenger ribonucleic acid; PIMMC=potential immune-mediated medical condition; SAE=serious adverse event; TEAE=treatment-emergent adverse event

**2019CoVn-313 Study Investigators**

| First name | Last name | Affiliation |
|---|---|---|
| Jeffrey | Adelglass | (Elite) Research Your Health |
| Adebayo | Akinsola | Tekton Research |
| Brandon | Alleman | Tekton Research |
| Codey | Bell | Tekton Research |
| Laurence | Chu | Benchmark Research |
| Matthew | Davis | Rochester Clinical Research |
| Sue | Fanning | AMR |
| David | Ferrera | Benchmark Research |
| George | Freeman | Health Research of Hampton Roads, Inc |
| Linda | Gorgos | (Elite) AXCES Research |
| Ripley | Hollister | (Elite) Lynn Institute of the Rockies |
| Michael | Jacobs | AMR |
| Craig | Julien | AMR |
| Karen | Kotloff | University of Maryland |
| Robert | Lockwood | Tekton Research |
| R. Scott | McClelland | University of Washington |
| Jara | McDonald | Tekton Research |
| Abel | Murillo | AMR |
| Robert | Noveck | AMR |
| Paul | Pickrell | Tekton Research |
| William | Seger | Benchmark Research |
| Stacy | Slechta | AMR |
| William | Smith | AMR |
| Harry | Studdard | AMR |
| Ronald | Surowitz | (Elite) Health Awareness, Inc. |
| Milagritos | Tapia | University of Maryland |
| Eduardo | Uribe | PanAmerican Clinical Research |
| Keith | Vrbicky | Velocity Clinical Research |
| Larkin | Wadsworth | Sundance Clinical Research, LLC |
| Kem | Yenal | (Elite) DM Clinical Research - Philadelphia |
| Pedro | Ylisastigui | AMR |
